# Estimation of the asymptomatic ratio of novel coronavirus infections (COVID-19)

**DOI:** 10.1101/2020.02.03.20020248

**Authors:** Hiroshi Nishiura, Tetsuro Kobayashi, Takeshi Miyama, Ayako Suzuki, Sung-mok Jung, Katsuma Hayashi, Ryo Kinoshita, Yichi Yang, Baoyin Yuan, Andrei R. Akhmetzhanov, Natalie M. Linton

## Abstract

A total of 565 Japanese citizens were evacuated from Wuhan, China to Japan. All passengers were screened for symptoms and also undertook reverse transcription polymerase chain reaction testing, identifying 5 asymptomatic and 7 symptomatic passengers testing positive for 2019-nCoV. We show that the screening result is suggestive of the asymptomatic ratio at 41.6%.

---

The number of novel coronavirus (COVID-19) cases worldwide continues to grow, and the gap between reports from China and statistical estimates of incidence based on cases diagnosed outside China indicates that a substantial number of cases are underdiagnosed (Nishiura et al., 2020a). Estimation of the asymptomatic ratio—the percentage of carriers with no symptoms—will improve understanding of COVID-19 transmission and the spectrum of disease it causes, providing insight into epidemic spread. Although the asymptomatic ratio is conventionally estimated using seroepidemiological data (Carrat et al., 2008; Hsieh et al., 2014), collection of these data requires significant logistical effort, time, and cost. Instead, we propose to estimate the asymptomatic ratio by using information on Japanese nationals that evacuated from Wuhan, China on chartered flights.

Figure 1 illustrates the flow of the evacuation process. By 6 February 2020 a total of *N*=565 citizens were evacuated. Among them, *pN*=63 (11.2%) were considered symptomatic upon arrival based on (1) temperature screening before disembarkation, and (2) face-to-face interviews eliciting information on symptoms including fever, cough, and other non-specific symptoms consistent with COVID-19. All passengers additionally undertook reverse transcription polymerase chain reaction (RT-PCR) testing, and *m*=5 asymptomatic and *n*=7 symptomatic passengers tested positive for 2019-nCoV.

**Figure 1.**
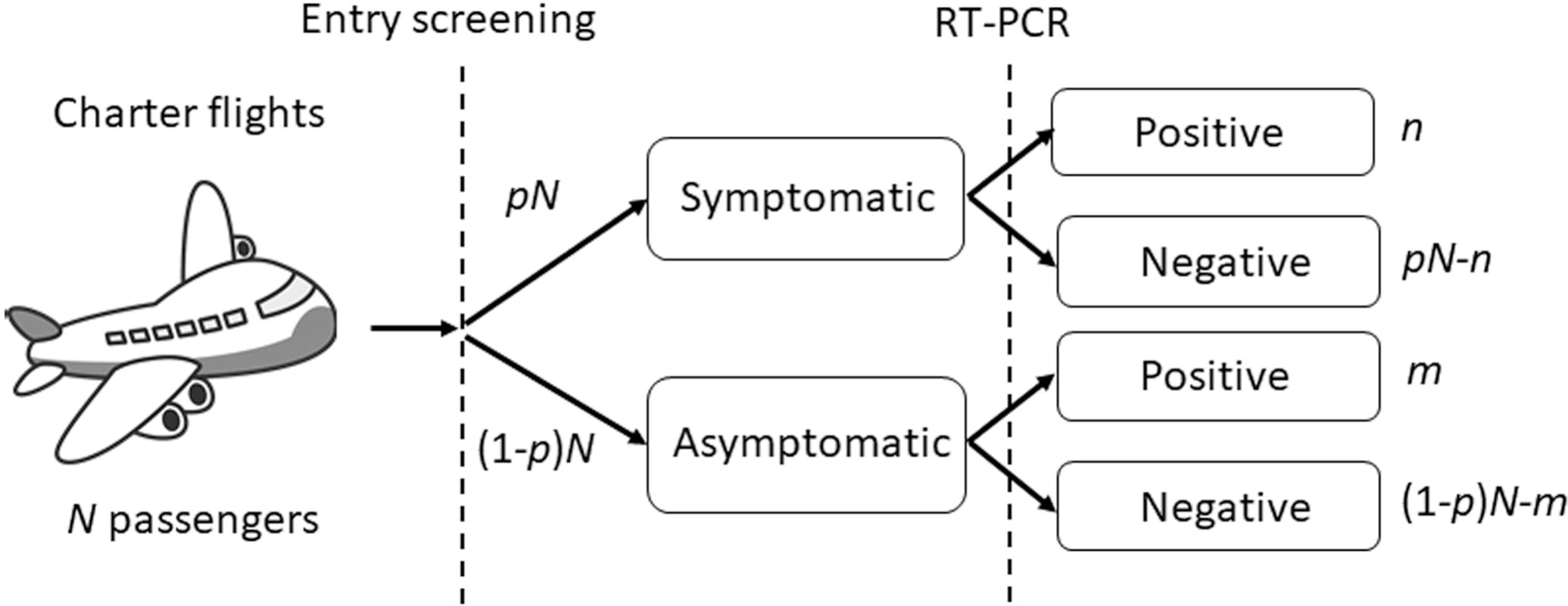
Flow of symptom screening and viral testing for passengers on chartered evacuation flights from Wuhan, China to Japan The flow of Japanese residents evacuating from Wuhan and screened in Japan. A total of *N* passengers were evaluated of which a fraction *p* were symptomatic upon arrival. Among symptomatic and asymptomatic individuals, *n* and *m* persons tested positive for the virus via reverse transcription polymerase chain reaction (RT-PCR).

Employing a Bayes theorem, the asymptomatic ratio is defined as

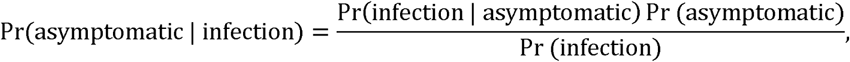

which can be calculated as *m*/(*n*+*m*), as seen in Figure 1. The asymptomatic ratio is thus estimated at 41.6% (95% confidence interval (CI): 16.7%, 66.7%) among evacuees.

Because fourteen days have elapsed since their departure from Wuhan— longer than the 95th percentile estimate of the COVID-19 incubation period (Li et al., 2020; Linton et al., 2020)—there is very little probability that the five virus-positive asymptomatic individuals will develop symptoms. Should one of the five becomes symptomatic in the future, the overall asymptomatic ratio would decrease to 33.3% (95% CI: 8.3%, 58.3%).

In general, asymptomatic infections cannot be recognized if they are not confirmed by RT-PCR or other laboratory testing, and symptomatic cases may not be detected if they do not seek medical attention (Nishiura et al., 2020b). Estimates such as this therefore provide important insight by using a targeted population to assess prevalence of asymptomatic viral shedding (Kupferschmidt & Cohen, 2020). Despite a small sample size, our estimation indicates that perhaps nearly a half of COVID-19-infected individuals are asymptomatic. This ratio is slightly smaller than that of influenza, which was estimated at 56–80% (Hsieh et al., 2014) using similar definitions for symptomatic individuals. There is great need for further studies on the prevalence of asymptomatic COVID-19 infections to guide epidemic control efforts.

## Data Availability

The data are available in the main text.

## Acknowledgments

H.N. received funding support from Japan Agency for Medical Research and Development [grant number: JP18fk0108050] the Japan Society for the Promotion of Science (JSPS) Grants-in-Aid for Scientific Research (KAKENHI in Japanese abbreviation) grant nos. 17H04701, 17H05808, 18H04895 and 19H01074, and the Japan Science and Technology Agency (JST) Core Research for Evolutional Science and Technology (CREST) program [grant number: JPMJCR1413]. NML received a graduate study scholarship from the Ministry of Education, Culture, Sports, Science and Technology, Japan. The funders had no role in study design, data collection and analysis, decision to publish, or preparation of the manuscript.

